# Intranasal oxytocin modulates the social salience network in adult men with autism

**DOI:** 10.64898/2026.08.24.26361211

**Authors:** Jacco G. Renström, Jelina Prinsen, Kaat Alaerts, Katrina Y. Choe

## Abstract

**Background:** Autism spectrum disorder is a prevalent neurodevelopmental condition featuring marked social difficulties. Oxytocin supplementation shows promising therapeutic efficacy in alleviating autism-like traits in rodent models, but clinical effects in humans remain inconsistent. The rodent-derived social salience network (SSN) comprises several oxytocin-modulated brain regions implicated in social behavior, but its conservation has not been established in humans. Here we assess, for the first time, functional connectivity (FC) within a homologous human SSN in autistic men to examine its relationship with behavioral traits and modulation by oxytocin.

**Methods:** The human SSN atlas was collated from open-access cortical and subcortical parcellations, and used to retrospectively analyze a resting-state fMRI dataset of adult men (N=28) with autism from a previously published, randomized, placebo-controlled oxytocin trial (1). SSN-wide and sub-network ROI-to-ROI FC correlations with social trait expression and salivary oxytocin concentrations were performed at baseline and post-administration. Treatment specific outcomes on FC were calculated using ANCOVA.

**Results:** We observed SSN sub-network FC correlations with social and repetitive behavioral scores (r≥0.401) and identified strong oxytocin sensitivity of nucleus accumbens–somatosensory and paraventricular nucleus–somatosensory circuits at baseline (r≥0.423). Following nasal spray administration, a strengthening of amygdala–somatosensory circuit was detected as the largest oxytocin-induced FC shift (q=0.04). Notably, baseline connectivity within this circuit strongly predicted treatment response (r=−0.651), with individuals having lower baseline FC showing greater post-treatment FC (q=0.033).

**Conclusions:** These findings provide first evidence for clinical relevance of the SSN in humans with autism and highlight circuits that may represent promising biomarkers for predicting oxytocin responsiveness.

## INTRODUCTION

Autism spectrum disorder is a prevalent neurodevelopmental condition commonly associated with difficulties in social communication and interaction, alongside restricted and repetitive behaviours (2,3). Critically, autism has a substantial and vastly multifaceted genetic etiology (4–6), posing a major challenge to the development of effective therapeutic support options for people with autism. To gain a mechanistic understanding of the relationship between individual genetic risk factors and the behavioral phenotypes, animal models of autism risk-gene mutations have been created and extensively characterized, many of which recapitulate core and co-occuring traits of autism (7,8). Convergent insights from several of these models have highlighted dysregulation of the neuropeptide oxytocin as a key contributor to the social behavioral phenotype in autism, including *Fmr1* (9) and *Nlgn1* (10), and *Cntnap2* (11,12) knockout animals. Moreover, oxytocin supplementation, either through endogenous activation or exogenous administration, successfully enhances social engagement in distinct genetic models of autism (10,13,14). Together, these preclinical observations provide a compelling mechanistic basis for pursuing oxytocin-based therapeutics as support options for autism.

However, despite this strong preclinical foundation and growing evidence linking endogenous oxytocin signalling to social traits in individuals with autism (15,16), translation of oxytocin therapies into human participants has been less conclusive. Whilst some trials report positive effects of intranasal oxytocin administration on several autism-related traits including social responsiveness and motivation (16–19), perceived attachment security (20), and socio-cognitive processing (21,22), as well as potential dose-dependet effects (23) other studies have failed to replicate these findings altogether (24–26). Furthermore, functional neuroimaging studies of oxytocin administration in autism have identified modulation of the amygdala as a key neural effect of treatment (Alaerts et al., 2019; Xiao et al., 2025); however, broader functional connectivity (FC) signatures relevant to apparent social traits remain insufficiently characterized and ambiguous (27,28). Given the substantial etiological heterogeneity of autism alongside the high dimensionality of neuroimaging data, where large-scale feature testing reduces statistical power through stringent multiple-comparison correction, constraining analyses to biologically defined networks with established relevance to oxytocin signalling and social behaviour may be recommended until sufficiently large datasets enable reliable whole-brain discovery (29).

One such candidate network is the putative social salience network (SSN) (30,31)—derived from rodent studies integrating socially-relevant sensory information to drive goal-directed social behaviour. Several regions within this network, including the paraventricular nucleus of the hypothalamus (PVN), nucleus accumbens (NAc), ventromedial prefrontal cortex (vmPFC), amygdala, and ventral tegmental area (VTA), are under oxytocinergic neuromodulatory input for regulating various aspects of social behaviour (31). Moreover, a recent large-scale multi-model functional neuroimaging study has highlighted dysregulation across a number of social behaviour-relevant regions including many SSN nodes (32). These findings are in accordance with our recent work showing oxytocin supplementation in *Cntnap2* knockout mice normalizes aberrant FC across SSN nodes along with elevating their level of social interaction (13). Furthermore, a growing body of research in autistic individuals has directly implicated circuit-level disruptions between key SSN regions, including the vmPFC, NAc and VTA (23,33–35). Collectively, these findings position the SSN and its modulation by oxytocin as a clinically relevant, cross-genetic point of convergence possibly underlying social traits in autism. However, the extent to which SSN FC is modulated by oxytocin administration in autistic individuals remain to be established.

In the present study we collated a human SSN atlas from open-source parcellations and retrospectively analyzed a previously published resting-state functional magnetic resonance imaging (rsfMRI) dataset of adult autistic men receiving single-dose intranasal oxytocin or placebo (1). The aims of our study were to examine (i) whether evolutionarily conserved oxytocin-sensitive SSN network signatures exist across mouse models and humans with autism, and (ii) whether SSN-focussed analysis can uncover previously undetected FC signatures associated with oxytocin signalling.

## METHODS

### Participants

This retrospective study analyzed rsfMRI data from 40 adult male individuals with an autism diagnosis (18-35 years old), receiving either oxytocin (n=22) or placebo (n=18) intranasally in a parallel, randomized, double blind trial; see Alaerts et al. (1) for detailed recruitment procedures. As the SSN includes critical parts of the reward circuity, such as the NAc and VTA, strongly modulated by dopaminergic medication for co-occurring symptoms, only unmedicated participants were included in this analysis, resulting in a final sample size of 28 (oxytocin n=14, placebo n=14). Additional follow-up analyses conducted in the full cohort to assess the robustness of the primary findings, and thereby further justify the sample selection, are presented in the *Supplemental Material*. Oxytocin (24 IU; Syntocinon; Sigma-Tau Industrie Farmaceutiche Riunite, Rome, Italy) or placebo (saline solution (0.9%) were self-administered intranasally using 15-mL glass bottles with metered pumps; ACA Pharma, Nazareth, Belgium). Written and informed consent was obtained from all participants at the time of the original study, and all procedures and data sharing were approved by Universitair Ziekenhuis/Katholieke Universiteit Leuven Ethics Committee for Biomedical Research (S56327) in accordance with The Code of Ethics of the World Medical Association (Declaration of Helsinki). The original trial was registered with the European Clinical Trial Registry (Eudract 2014-000586-45) and the Belgian Federal Agency for Medicines and Health Products.

### Measures

Demographic measures, including Age, IQ and Autism Diagnostic Observation Schedule (36) scores used as a diagnostic tool for autism presentation are shown in Table 1. Behavioral assessments used for statistical comparisons included the Social Responsiveness Scale (SRS) (37), which measures expression of key autism traits within the domains of social communication, social awareness, social motivation, and restricted/repetitive behaviors; the Repetitive Behavior Scale–Revised (RBS) (38), which assesses the frequency and severity of repetitive behaviors; the State Adult Attachment Measure (SAAM) (39), which evaluates attachment security, anxiety, and avoidance. Salivary oxytocin was used as a non-invasive index of peripheral oxytocin concentrations; however, its correspondence with central oxytocinergic activity remains uncertain and should be interpreted cautiously (40,41). Salivary samples were collected immediately preceding oxytocin administration using cotton swabs, and oxytocin concentrations were measured using a commercially available enzyme-linked immunosorbent assay following standard collection and processing procedures. Anatomical (repetition time = 9.4 ms, echo time = 4.6 ms, 1.2 mm^3^ voxel size) and subsequent 7-min rsfMRI scans (gradient-echo planar imaging sequence, repetition time = 2500 ms, echo time = 30 ms, 2.5 mm^3^ voxel size) covering the entire brain and brainstem were collected at baseline and 30 min post-treatment administration using a 3T magnetic resonance scanner (Phillips Healthcare, Best, the Netherlands).

**Table 1.** Demographic and clinical characteristics of participants included in retrospective analysis. Group means were compared across each score using student *t* tests.

|  | **Oxytocin (n=14)** |  | **Placebo (n=14)** |  | **p** |
| --- | --- | --- | --- | --- | --- |
| Age (Years, mean±SD) | 23.29 | 3.97 | 23.57 | 5.68 | 0.88 |
| **IQ, mean±SD** |  |  |  |  |  |
| Total IQ | 99.64 | 12.23 | 107.71 | 19.66 | 0.20 |
| Verbal IQ | 104.79 | 8.93 | 112.14 | 13.88 | 0.11 |
| Performance IQ | 102.86 | 19.11 | 103.50 | 22.77 | 0.94 |
| **ADOS Score, mean±SD** |  |  |  |  |  |
| Total | 7.14 | 4.69 | 8.36 | 3.43 | 0.44 |
| Communication | 2.14 | 1.10 | 2.50 | 1.40 | 0.46 |
| Social interaction | 5.00 | 3.86 | 5.86 | 2.93 | 0.51 |
| Stereotypical behavior | 1.14 | 1.41 | 1.07 | 0.92 | 0.88 |

### Data Preprocessing and analysis

Prior to data sharing, all scans were defaced at KU Leuven using PyDeface (v2.0.2) within FSL, and header information was removed using the MRtrix3 *mrconvert* function. Subsequent data preprocessing was performed at McMaster University, following closely the reported steps in the previous publication. rsfMRI scans were preprocessed and analyzed using the CONN functional connectivity toolbox 22.v2407 (42) implemented in MATLAB R2024b (Mathworks). Preprocessing steps included realignment and normalization to standard space, resampling (3-mm isotropic), confound regression (CompCor (43)), scrubbing (framewise displacement > 0.5 mm) and bandpass filtering (0.009 < f < 0.08Hz). For ROI-ROI analysis, SSN ROIs (n=8) (Figure 1A) were collated using inbuilt Harvard-Oxford Cortical Structural Atlas (RRID:SCR_001476) (44). The ROI label indices were as follows: somatosensory cortex [S1] (labels: 33 & 34), auditory cortex (labels: 82 & 83), visual cortex [V1] (labels: 90 & 91), amygdala (labels: 102 & 103), nucleus accumbens (labels: 104 & 105). As the vmPFC is a functional collection of prefrontal sub-regions it was combined using ROIs for the frontal pole (ventral part) (labels: 1 & 2), frontal medial cortex (label: 49), subcallosal cortex (label: 52) and anterior cingulate gyrus (label: 55). Finally, subcortical parcellations of the PVN and VTA were added using open-source parcellations that have been previously validated on 3T fMRI data (45–47). It should be noted that, due to the inherent size of the PVN parcellation in combination with smoothing, PVN signal likely reflects general activity within the wider ventromedial hypothalamus. However, given its central role within the SSN it was included in the analysis.

**Figure 1.**
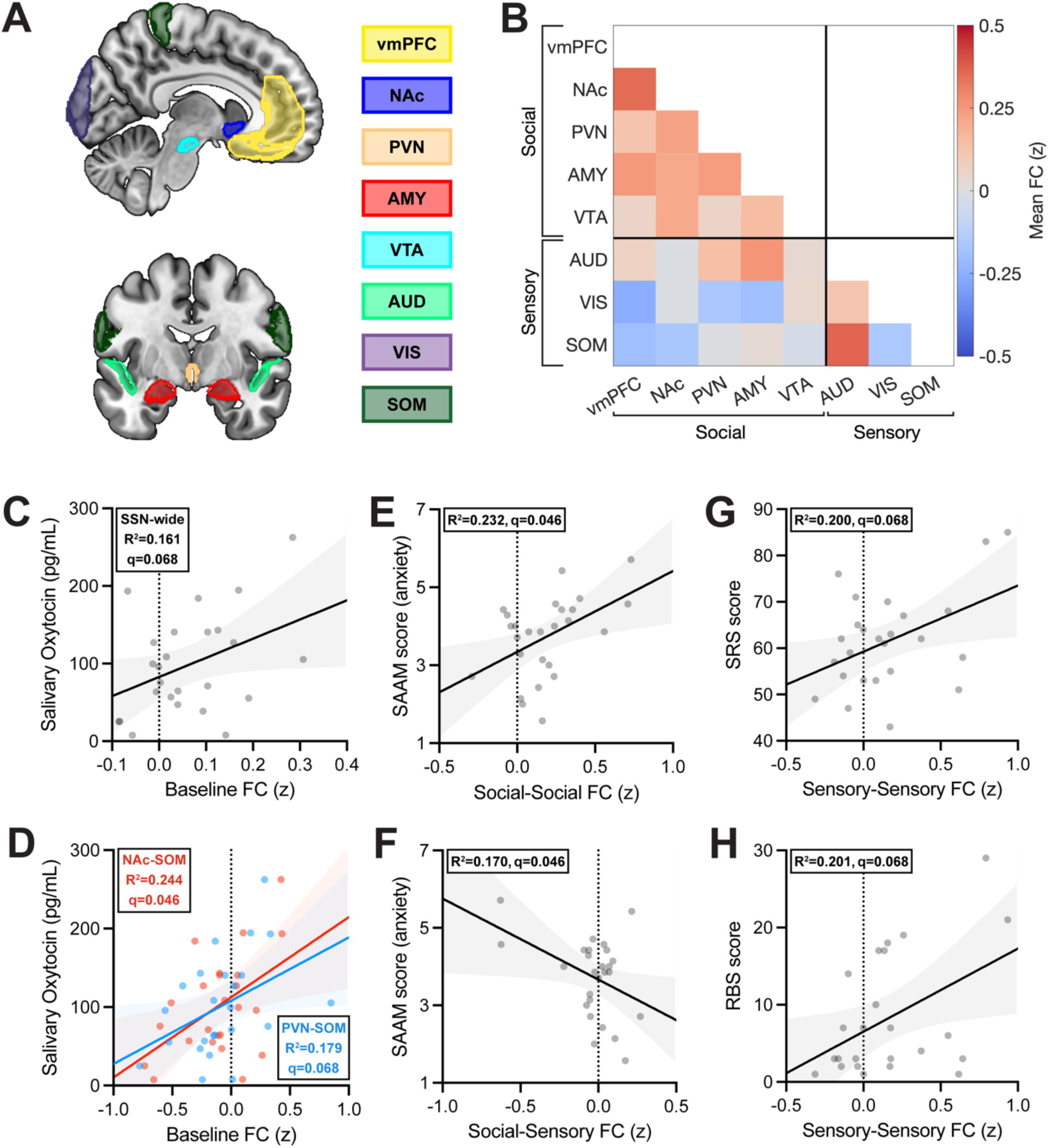
Baseline SSN-wide and sub-network FC correlate with social trait expression and circulating oxytocin concentrations. A) SSN ROI masks used in the retrospective analysis projected onto a standard MNI152 brain template. vmPFC-ventromedial prefrontal cortex (yellow), NAc-Nucleus Accumbens (dark blue), PVN-paraventricular nucleus (light brown), AMY-amygdala (red), AUD-auditory cortex (light green), VIS-visual cortex (purple), VTA-ventral tegmental area (light blue), PVN-paraventricular nucleus (light brown), SOM-somatosensory cortex (dark green). B) Baseline z-scored FC across SSN ROIs, with Social/Sensory categorization. Warm to cool shading representing greater positive to negative FC, respectively. C) Correlation between basal SSN-wide FC averaged across all edges and salivary oxytocin concentration. D) Correlation between basal NAc-SOM FC (red) or PVN-SOM FC (blue) and salivary oxytocin. E-H) Scatterplots showing correlations between basal inter- and intra-domain FC (z-scored) and social trait scores. Across all scatterplots, individual datapoints shown, solid lines represent fitted linear regression line with shaded region around regression line indicating 95% confidence intervals. Linear fit and familywise corrected p-value (FDR<0.05) for plot/group shown in upper hand corner of each graph.

Mean FC values were calculated by averaging pairwise FC across either the entire SSN or within each sub-network (social-social, social-sensory, and sensory-sensory). Given the widely documented placebo effect in fMRI studies (19,48,49) a two-stage thresholding procedure was applied to identify the strongest oxytocin-specific connections prior to statistical comparison. First, edges were baseline-normalized and then ranked according to the magnitude of treatment-related change, calculated as ΔOxytocin − ΔPlacebo. Then edges were thresholded so only edges exhibiting greater change in the oxytocin group than the placebo group (|ΔOxytocin| > |ΔPlacebo|) were retained. Outlier removal (ROUT, Q=1%) and subsequent data analysis was carried out using GraphPad Prism 10. Familywise p-values were corrected for multiple comparisons using the Benjamini–Hochberg false discovery rate procedure (FDR, q<0.05).

## RESULTS

### Oxytocinergic and behavioral signatures of the SSN in autism

First, we examined one of the proposed defining properties of the SSN—its sensitivity to oxytocin (31,50). Because several key subcortical SSN nodes are directly modulated by psychotropic medications, only unmedicated adults were included in this analysis. Baseline rsfMRI scans from our placebo and oxytocin participant groups (acquired prior to nasal spray administration) were combined (n=28) to examine associations between SSN-wide FC and oxytocin levels (Figure 1B). We observed a strong correlation between mean SSN FC and basal salivary oxytocin concentration; however, this did not survive correction (r=0.401, q=0.068, 95% CI [0.01, 0.68], Figure 1C). As a result, we examined individual pairwise ROI FC and salivary oxytocin levels at baseline, which revealed a strongest association between endogenous oxytocin and NAc- somatosensory FC (r=0.494, q=0.046, 95% CI [−0.11, 0.75], Figure 1D). Additionally, a near-threshold association between PVN-somatosensory FC was also observed, (r=0.423, q=0.068, 95% CI [−0.02, 0.71]; next largest |r|=0.277, q=0.205). These results indicate that, despite the smaller sample size and ongoing debate regarding salivary oxytocin as a proxy for central oxytocin activity (see Martins et al. (41)), individuals with higher salivary oxytocin levels exhibit stronger integration within the SSN. Behaviorally, the strong association between basal oxytocin levels and SAAM security scores, highlighted in our previous report of the same sample (1), was also observed here (r=0.676, q=0.013, 95% CI [0.38, 0.85]; not shown), indicating preservation of key behavioral correlates despite the smaller group sizes.

Next, associations between SSN-wide FC and behavioral measures relevant to autism were explored. At the network level, there were no significant correlations with any measures, albeit mean SSN-wide FC showed near-threshold positive correlations with total SRS score (r=0.367, q=0.085, 95% CI [−0.24, 0.66]) and RBS score (r=0.356, p=0.100, 95% CI [−0.05, 0.66]) (next largest |r|=0.220, q=0.280).

Given this lack of network-wide association with behavioral trait expression, we hypothesized that behavioral traits are better represented by FC within functional sub-networks of SSN. To test this, the SSN was divided into two sub-networks based on their established functional specialization: ‘social’ or ‘sensory’ (Figure 1B). Sensory regions included the auditory, visual, and somatosensory cortices, whereas social regions comprised the vmPFC, NAc, PVN, amygdala, and VTA. In line with our hypothesis, we found specific sub-network FC better represented individual behavioural measures: FC among social SSN regions was positively associated with SAAM attachment anxiety scores (r=0.482, q=0.046, 95% CI [0.12, 0.73]) (Figure 1E), whereas FC between social and sensory SSN regions was negatively correlated with SAAM attachment anxiety (r=−0.412, q=0.046, 95% CI [−0.69, −0.03]) (Figure 1F). Additionally, we observed a near-threshold positive association between FC among sensory SSN nodes and SRS and RBS scores (r=0.401, 95% CI [0.06, 0.72], Figure 1G; r=0.414, 95% CI [0.05, 0.72], Figure 1H, respectively; both q=0.068).

### Oxytocin-responsive pathways within the SSN

Next, we explored whether intranasal oxytocin modulates FC across the SSN. Given the established role of oxytocin signalling across several key SSN nodes (50), we first examined its effects on SSN FC at the network level. Baseline and post-treatment FC in placebo and oxytocin groups were compared using a repeated-measures ANOVA (Figure 2A). No main effect of treatment was detected for SSN-wide FC or any sub-network FC (all F<1) (Figures 2B-E). Together, these findings indicate that SSN FC is associated with both behavioral measures and endogenous oxytocin levels in autism, while intranasal oxytocin does not appear to globally alter SSN FC.

**Figure 2.**
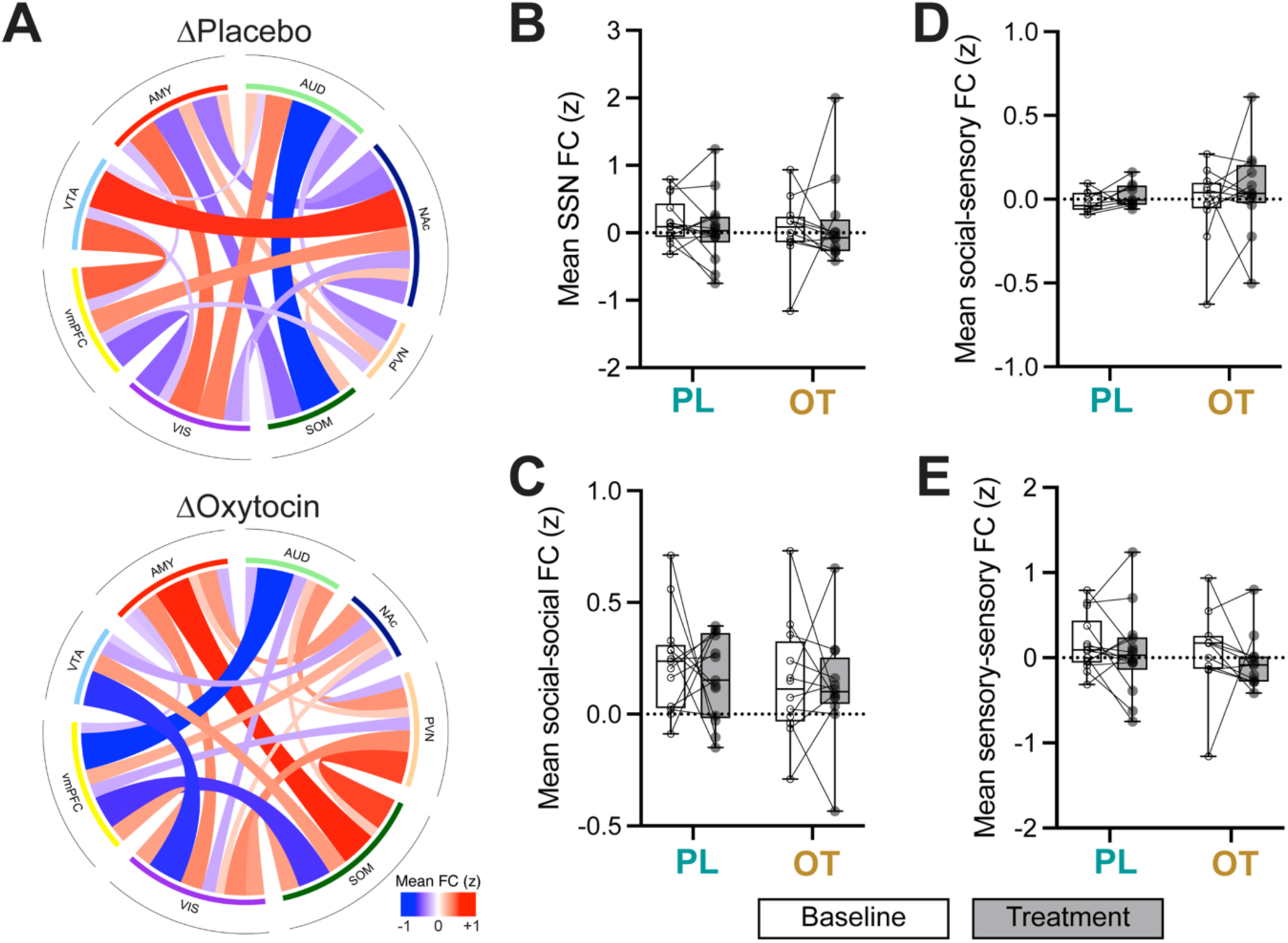
Oxytocin does not modulate FC at the network- or sub-network levels. 2A) Placebo (top panel) and oxytocin (bottom panel) delta FC changes (baseline – post-treatment) across entire SSN. Cool-to-warm colours indicate reduced-to-increased FC in response to treatment. B-E) Bar plots showing the mean FC change across baseline (white) and post-treatment (grey) between groups (PL-placebo, OT-oxytocin). Individual subjects shown as points with connected lines. Data was analysed using a 2-way repeated measures ANOVA with FDR-correction<0.05 but did not reveal any significant changes.

We next examined the effects of oxytocin on individual connections within the SSN. Following our two-stage thresholding procedure detailed above, the resulting 11 edges represented connections between SSN nodes exhibiting greater FC changes in the oxytocin group compared to placebo, explaining 46.14% of the total treatment variance related to FC change (Figure 3A): amygdala- somatosensory (Δ_O-P_=0.95), vmPFC-somatosensory (Δ_O-P_=−0.51), vmPFC-auditory (Δ_O-P_=−0.50), PVN-auditory (Δ_O-P_=0.49), and PVN-somatosensory (Δ_O-P_=0.44), visual-VTA (Δ_O-P_=−0.40), somatosensory-VTA (Δ_O-P_=0.40), somatosensory-visual (Δ_O-P_=0.40), auditory-amygdala (Δ_O- P_=−0.35), visual-PVN (Δ_O-P_=0.30), and VTA-amygdala (Δ_O-P_=−0.10). Among these 11 edges, a statistically significant interaction between treatment and time (baseline/post-treatment) was detected for amygdala-somatosensory FC (F(1,20)=7.57, p=0.01; two-way repeated-measures ANOVA) (Figure 3B). Post-hoc analyses revealed greater FC following oxytocin administration compared to baseline (q=0.004), but not placebo (q=0.325). Specifically, oxytocin appeared to normalize this connection from negative FC at baseline (mean z=−0.44) to positive FC following treatment (mean z=0.49). No additional significant effects were detected across all examined FC changes (all F<1), but it is worthwhile to mention that PVN–somatosensory FC approached our significance threshold (F(1,23)=3.92, p=0.06) (Figure 3C).

**Figure 3.**
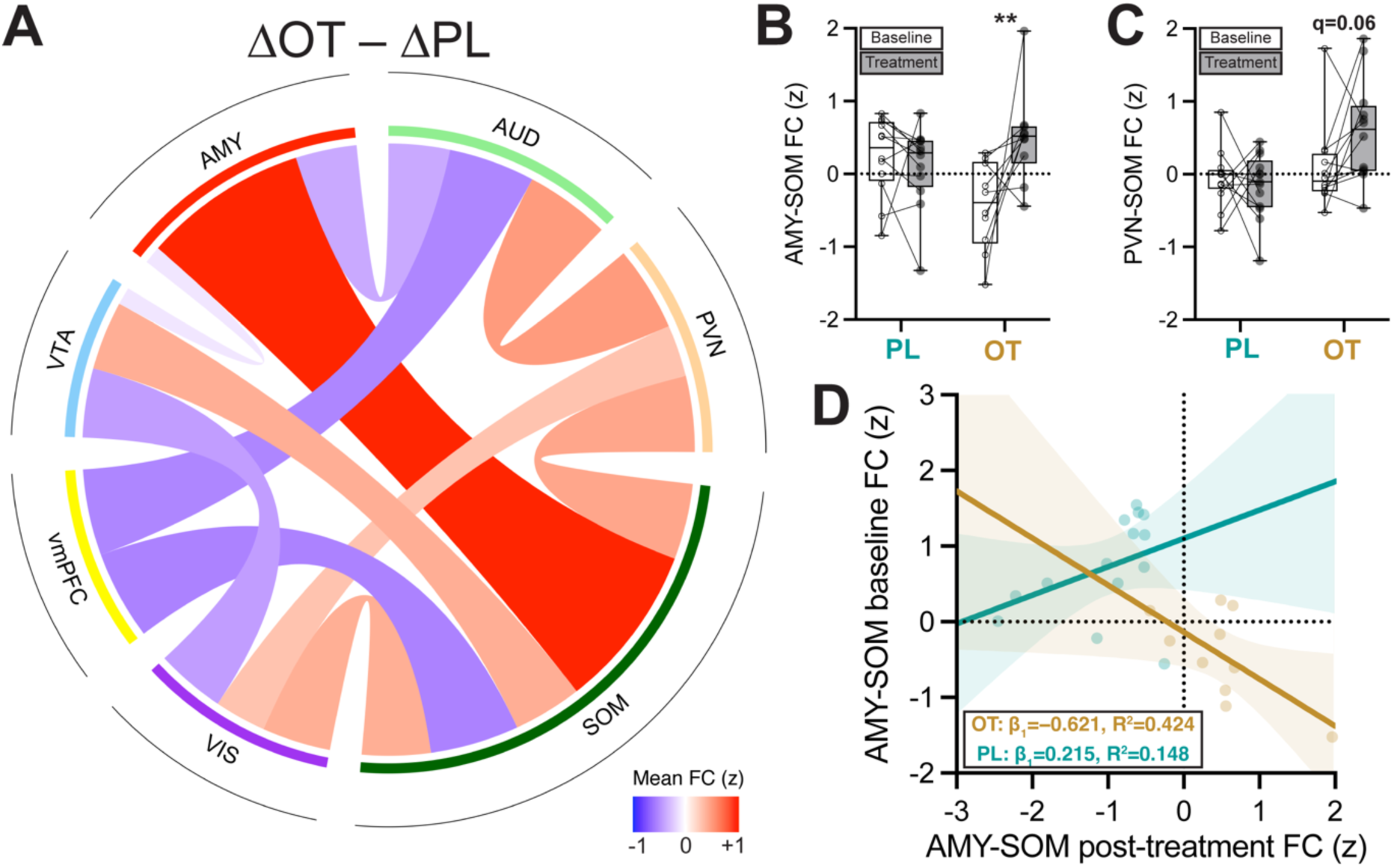
Oxytocin robustly modulates amygdala-somatosensory FC, with a dependency on basal connectivity. A) Chord plot depicting oxytocin-specific change in FC across SSN (ΔOxytocin – ΔPlacebo). Warmer colours indicate greater positive DFC post oxytocin-treatment. Line width indicates magnitude of change, irrespective of direction. B-C) Box plots highlighting two edges with the largest effect size post-analysis: amygdala-somatosensory (AMY-SOM) and PVN-somatosensory (PVN-SOM). Mean FC (z-scored) shown at baseline (white) and post-treatment (grey). Individual subjects shown as points with connected lines. Data was analysed using a 2-way repeated measures ANOVA, with FDR-correction<0.05: **=q=0.004. D) Scatter plot with fitted linear regression lines (solid line) and 95% confidence intervals (shaded regions) for each condition (Oxytocin – gold, Placebo – turquoise) for AMY-SOM FC at baseline and post-treatment. Slope and R^2^ of the linear regression shown for each group shown in lower hand corner of the graph (OT – oxytocin, PL – placebo).

Despite evidence of individual differences in basal connectivity and endogenous oxytocin concentrations predicting longitudinal outcomes in autism (16,51), no reliable functional connectivity biomarker of pharmacological efficacy has been identified to-date. Given the robust modulation of amygdala-somatosensory FC observed here, we further explored whether baseline FC could be used as a candidate predictor for post-treatment FC. To assess whether baseline connectivity differentially predicted post-treatment connectivity as a function of treatment, an ANCOVA was performed with post-treatment amygdala-somatosensory FC as the dependent variable, treatment (oxytocin vs placebo) as the fixed factor, and baseline amygdala- somatosensory FC as a covariate (Figure 3D). This revealed a significant treatment x baseline

FC interaction (F(1,22)=5.15, p=0.033, uncorrected), indicating that the relationship between baseline and post-treatment connectivity differed between the oxytocin and placebo groups. Inspection of the fitted regression lines showed a more pronounced, negative slope in the oxytocin group (β_1_=−0.621, R^2^=0.424), compared to the placebo group (β_1_=0.215,R^2^=0.148), indicating lower baseline amygdala-somatosensory FC is more greatly modulated following oxytocin, but not following placebo administration.

Given this strong predictive relationship between baseline amygdala-somatosensory FC and oxytocin-induced FC changes, we examined whether the baseline FC for this edge is a relevant parameter for any autism-related behavioural domains assessed in this cohort. We found baseline amygdala-somatosensory FC was not correlated with any behavioral scores or oxytocin levels measured prior to treatment (largest |r|=0.273, q=-408), suggesting that the amygdala- somatosensory circuit may be a predictive biomarker of oxytocin responsiveness, rather than a biomarker of clinical trait expression.

## DISCUSSION

In this retrospective analysis of autistic adult men, we investigated whether the SSN, an oxytocin-modulated brain network constructed based on rodent experimental data, exhibits evidence of conservation and clinical relevance in humans. Two principal findings emerged. First, we found that the putative SSN displays functional conservation, oxytocin sensitivity, and clinical relevance in human participants with autism. By partitioning the network into social and sensory sub- networks and assessing connectivity, we found that interactions within and between these domains may contribute to several autism-relevant behavioural features, including repetitive behaviours and social attachment-related traits. Second, intranasal oxytocin did not induce global alterations in SSN FC, but instead selectively modulated a restricted subset of network connections, with amygdala-somatosensory FC emerging as the most robust oxytocin-sensitive pathway. Together, these findings provide novel evidence that the putative SSN we present here captures biologically and behaviourally meaningful variations in autism, and may represent a translational framework linking oxytocin signalling, socio-sensory processing, and individual variability in treatment response.

### Evidence for cross-species conservation and clinical relevance of the SSN

The present findings provide converging support for translational conservation of the SSN across species, which we assessed along two key dimensions: (i) whether reduced SSN connectivity signatures were associated with increased social or sensory phenotypes, and (ii) whether these SSN signatures correlated with endogenous oxytocin concentrations in a congruent manner. With respect to (i), we found that a hyperconnectivity within sensory SSN regions tracked greater autism trait expression, while disrupted coupling within social SSN regions and between sensory– social SSN components were linked to increased attachment-related anxiety (Fig. 1E-F). These observations are consistent with contemporary models proposing atypical sensory processing representing a fundamental contributor to social difficulties in autism rather than a secondary consequence of reduced social interaction (23,52).

In support of (ii), we observed SSN-wide and circuit-level FC were positively associated with salivary oxytocin concentrations (Fig. 1G-H). This relationship is consistent with the proposed neurochemical basis of the SSN, as oxytocin receptors are densely expressed across several key SSN nodes, including the amygdala, nucleus accumbens, hypothalamus, and VTA (50), and oxytocin signalling is critically involved in modulating the salience and processing of social stimuli (53). Thus, intrinsic SSN FC may provide a systems-level readout of endogenous oxytocinergic function, capturing individual variability in a neurochemical pathway directly implicated in social behaviour. This is particularly relevant in autism, where substantial heterogeneity in endogenous oxytocin concentrations has been reported (54), potentially contributing to variability in oxytocin treatment responsiveness. Consistent with this possibility, individuals with autism exhibiting lower endogenous oxytocin levels show the greatest improvements in SRS scores following oxytocin administration (16). Together, these findings identify SSN FC as a candidate biomarker for characterizing endogenous oxytocin system function and potentially stratifying individuals who may be more likely to benefit from oxytocin-based interventions.

However, because behavioral measures were collected only prior to oxytocin administration, we were unable to determine whether treatment-induced changes in SSN FC were associated with subsequent improvements in social behaviour. Moreover, measures such as the SRS and SAAM primarily capture relatively stable social traits and attachment-related characteristics and may therefore be insensitive to acute neurobehavioral effects of oxytocin (39,55). Future longitudinal studies incorporating chronic oxytocin administration and repeated behavioral assessments will therefore be necessary to determine whether oxytocin-induced changes in SSN FC translate into notable behavioral improvements.

### Selective oxytocin modulation of amygdala-somatosensory circuitry

Despite our results linking SSN FC to endogenous oxytocin signalling (Fig 1C&D), acute oxytocin administration did not alter global network FC (Fig. 2B). Instead, we show that oxytocin appears to exert selective effects on specific circuits (Fig. 3A-C). Indeed, the strongest treatment effect of oxytocin was observed in the amygdala-somatosensory pathway (Fig. 3D). The involvement of the amygdala in this subsampled dataset is consistent with what we have previously reported using the full dataset (1), and also previous literature highlighting the amygdala as one of the most reliable neural targets of oxytocin administration in autism (28), often reducing threat-related responses while enhancing the processing of socially relevant information (56–59). Contrastingly, our finding of somatosensory cortex involvement in two of the largest oxytocin-induced FC shifts within the SSN (i.e. amygdala-somatosensory cortex and PVN-somatosensory cortex) is somewhat unexpected. The somatosensory cortex has classically been ascribed a more peripheral, predominantly affiliative touch-related, role in social cognition (60). However, growing evidence from both animal model and human studies suggests this cortical area and its modulation by oxytocin may be more integral to broader social cognition than previously appreciated. For example, Zheng et al. (61) demonstrated that early-life sensory deprivation in mice, via whisker trimming, reduces excitatory synaptic transmission in the somatosensory cortex and disrupts development of the central oxytocin system. Furthermore, acute oxytocin infusion into the somatosensory cortex of these mice was sufficient to rescue the excitatory synaptic phenotype. Subsequent work extended these findings by linking early sensory perturbations to persistent social and anxiety-related behavioural phenotypes in adulthood, including in autism-risk gene models (62,63). In humans, neuroimaging studies have also indicated that cortical somatosensory regions contribute to subcortical processes (e.g., within the amygdala) in processes of emotion recognition, social perception, empathy, and the representation of affective states in both self and others (64,65), whilst altered sensory responsivity has been linked to social difficulties, repetitive behaviours, and anxiety across the autism spectrum (52). Within the context of autism, these observations suggest its social symptoms may arise not solely from functional differences within brain regions with specifically ascribed functions in social behaviour, but perhaps also from broader disrupted socio-sensory circuitry (66). Thus, our observations prompt further investigation in autistic and non-autistic individuals to examine how social and sensory information are integrated within the brain to modify social behaviour, and how oxytocin modulates this process.

Interestingly, we observe baseline amygdala-somatosensory FC strongly predicted post-treatment connectivity, with oxytocin producing marked normalization of the baseline hypoconnectivity phenotype. These robust findings suggest that amygdala-somatosensory connectivity may represent both a functionally relevant target of oxytocin within the SSN and a potential source of inter-individual variability in treatment response. Mechanistically, the coordinated activity between the amygdala and somatosensory cortex may arise through intermediary hubs involved in salience attribution and multisensory integration rather than a monosynaptic connection (67). In support of this, Guo and colleagues (68) observed autism-specific hypoconnectivity between the amygdala and thalamus, a key hub for somatosensory integration, which correlated significantly with social trait expression. Given the established role of the amygdala in assigning affective significance to biologically relevant stimuli (69,70), simultaneous recruitment of sensory and affective systems under the mediation of oxytocin may provide a mechanism through which socially salient sensory experiences are selectively reinforced and consolidated. Such a framework is consistent with evidence that oxytocin promotes synaptic and circuit plasticity across distributed brain regions (61,71), and with contemporary theories proposing that oxytocin acts primarily as a facilitator of social learning and salience-dependent information processing rather than as a universally prosocial signal (53,72). Notably, despite the robust neural effect observed, we did not identify significant behavioural correlates of baseline amygdala-somatosensory connectivity, and it remains difficult to assess clinical-behavioral changes upon single-dose pharmacotherapy. Consequently, any link between modulation of amygdala-somatosensory FC and behavioural outcomes remain speculative and should be examined in a larger cohort study.

## Conclusion

In summary, the present study provides the first demonstration of the putative SSN in humans that potentially represents behavioral and oxytocin-related signatures in humans with autism paralleling observations from animal models. Our findings support emerging precision-medicine approaches based on the hypothesis that oxytocin may be most effective in biologically defined subgroups rather than across autism populations as a whole. Our results also identify amygdala- somatosensory FC as a promising candidate biomarker for oxytocin treatment stratification. Several limitations should be acknowledged, including the modest sample size, restriction to unmedicated adult men, the exploratory nature of the edge-selection procedure, and the absence of longitudinal behavioral outcome measures. Nevertheless, by linking oxytocin signaling, socio- sensory processing, and individual variability in treatment response, this work positions the SSN as a potentially conserved translational framework for understanding social differences in autism and an intriguing prospective biomarker for oxytocin treatment efficacy.

## Data Availability

All data produced in the present study are available upon reasonable request to the authors.

## Supplemental material

### Effect of Including Participants Receiving Psychotropic Medication

To assess the robustness of the primary findings, all analyses were repeated in the full cohort, including participants receiving psychotropic medication (N=36). Overall, inclusion of medicated participants did not substantially alter the principal findings but did provide additional justification for their exclusion on the current report. Baseline SSN-wide functional connectivity and PVN–somatosensory connectivity remained near-threshold association with basal salivary oxytocin concentrations (SSN-wide: r=0.372, q=0.071, 95% CI [0.03, 0.63]; PVN–somatosensory: r=0.353, q=0.071, 95% CI [0.01, 0.63]), whereas the association between NAc–somatosensory connectivity and salivary oxytocin did not survive FDR correction (r=0.283, q=0.116, 95% CI [−0.07, 0.58]). This attenuation may reflect medication-related differences in NAc function. Indeed, we observed a near-threshold baseline difference in NAc-somatosensory FC (ANOVA: F(1,33)=3.29, q=0.079) tending to exhibit greater NAc-somatosensory FC compared to unmedicated participants. Moreover, exploration of the medication status was significantly associated with both Social Responsiveness Scale (SRS) scores (F(1,34)=4.93, q=0.033) and Repetitive Behavior Scale (RBS) scores (F(1,33)=8.14, q=0.014). Together, these findings suggest that psychotropic medication may contribute to variability in both clinical phenotype and specific aspects of SSN functional organisation, underscoring the importance of medication-based stratification in the current and future investigations of SSN function and oxytocin-related neural mechanisms in autism.

Notably, however, the treatment-related increase in amygdala–somatosensory connectivity observed here remained significant after including medicated participants in the ANCOVA (F(1,32)=4.35, p=0.045, uncorrected), indicating that the principal neural effect of intranasal oxytocin was robust to medication status.

## Acknowledgements

This dataset was previously published (1), and was kindly made available for further re-analysis by Dr. Alaerts and Dr. Prinsen. Additionally, we thank Dr. Bern Accou (KU Leuven) for facilitating the sharing of this dataset. We also extend special thanks to Sylvie Bernaerts, supported by a Marguerite-Marie Delacroix doctoral fellowship, for her central role in collecting the data analysed in this study. Collection of the original dataset was further supported by the Branco Weiss Fellowship of the Society in Science, Eidgenössische Technische Hochschule Zurich (to KA), Flanders Fund for Scientific Research Grant Nos. KAN 1506716N, KAN 1521313N, and G.0401.12 (to KA). Additionally, KYC was supported by the Canada Research Chair Program Tier 2 (CRC-2020-00071), CIHR project grant (PJT-183808). KA obtained supported via the Research Foundation – Flanders (FWO), Scientific Research Network (W000425N) and KU Leuven Research Council, Global Seed Fund 2026 (GSF26065).

## Declaration of generative AI and AI-assisted technologies in the writing process

During the preparation of this work the authors used ChatGPT 5-5 in order to refine text and improve readability. After using this tool, the authors reviewed and edited the content as needed and take full responsibility for the content of the publication.

